# Paradox of Predictors in Critically ill COVID-19 Patients: Outcome of a COVID-dedicated Intensive Care Unit

**DOI:** 10.1101/2021.04.23.21256009

**Authors:** Morshed Nasir, Rawshan Ara Perveen, Rumana Nazneen, Tahmina Zahan, Sonia Nasreen Ahmad, ASM Salauddin Chowdhury

**Affiliations:** Holy Family Red Crescent Medical College, Dhaka, Bangladesh; Holy Family Red Crescent Medical College Hospital, Dhaka, Bangladesh

**Keywords:** COVID-19, critical care, comorbidities, intensive care unit, biomarkers, mortality

## Abstract

**Background:** The study aimed to analyze the demographic, comorbidities, biomarkers, pharmacotherapy, and ICU-stay with the mortality outcome of COVID-19 patients admitted in the intensive care unit of a tertiary care hospital in a low-middle income country, Bangladesh.

**Methods:** The retrospective cohort study was done in Holy Family Red Crescent Medical College Hospital from May to September 2020. All 112 patients who were admitted to ICU as COVID-19 cases (confirmed by RT-PCR of the nasopharyngeal swab) were included in the study. Demographic data, laboratory reports of predictive biomarkers, treatment schedule, and duration of ICU-stay of 99 patients were available and obtained from hospital records (non-electronic) and treatment sheets, and compared between the survived and deceased patients.

**Results:** Out of 99 patients admitted in ICU with COVID-19, 72 were male and 27 were female. The mean age was 61.08 years. Most of the ICU patients were in the 60 - 69 years of age group and the highest mortality rates (35.89%) were observed in this age range. Diabetes mellitus and hypertension were the predominant comorbidities in the deceased group of patients. A significant difference was observed in neutrophil count, creatinine and, NLR, d-NLR levels that raised in deceased patients. There was no significant difference as a survival outcome of antiviral drugs remdesivir or favipiravir, while the use of cephalosporin was found much higher in the survived group than the deceased group (46.66% vs 20.51%) in ICU.

**Conclusions:** Susceptibility to developing critical illness due to COVID-19 was found more in comorbid males aged more than 60 years. There were wide variations of the biomarkers in critical COVID-19 patients in a different population, which put the healthcare workers into far more challenge to minimize the mortality in ICU in Bangladesh and around the globe during the peak of the pandemic.

## Introduction

The coronavirus disease COVID-19 pandemic is the defining global health crisis at present and the greatest challenge since World War Two. The public healthcare system is suffering an unprecedented challenge due to the COVID-19 pandemic on a global scale. Globally 217 countries have been affected with Novel coronavirus disease 2019 (COVID-19) disease till now. COVID-19, now regarded as a pandemic by the World Health Organization. The cumulative numbers to over 79 million reported cases and over 1.7 million deaths globally since the start of the pandemic^1^. This huge number of cases infected with the new SARS-CoV-2 virus demonstrates the highly contagiousness that outstripped any public health disaster caused by any single disease in human history^2^. Infection rates and deaths throughout the world have been raised exponentially. In China and Italy, initially, mortality ranging from 26% to 62% in critically ill patients. Whereas, in Seattle and New York, the mortality range varied from 23% to 50% respectively^3^.

In the case of 80% of patients, SARS-COV-2 infections are self-limiting and special treatment may not require,15% of the infected patients may present with co-morbidities such as diabetes mellitus, ischemic heart disease, hypertension, and obesity are more likely to develop severe pneumonia, get admitted to the health facilities with disease progression to get proper care and rest 5% progress to respiratory failure, acute respiratory distress syndrome (ARDS) and need intensive care unit (ICU) support for a long period of time^4^. Older age and co-morbid disease have been reported as risk factors for death.

Severe inflammatory and immunological response caused by infectious disease, and its significant role in the progression of viral pneumonia, including COVID-19 that may lead to weak adaptive immune response and imbalance^5^. Therefore, the circulating biomarkers that may represent the inflammatory and immune response are vigorously studied as a potential predictor for the prognosis of the disease. But there are unexplained variations in different populations, severity, and demography. Considering the useful predictors for the prognosis of patients with viral pneumonia, some hematological count like white blood cell (WBC) count, neutrophil(N), lymphocyte (L), neutrophil-lymphocyte ratio (NLR), neutrophil count divided by the result of WBC count minus neutrophil count (d-NLR), platelet, platelet-to-lymphocyte ratio (PLR) are investigated. Some inflammatory biomarkers like ferritin, d-Dimer, C-reactive protein (CRP) are also monitored at different severity of disease^6^.

Patients with severe COVID-19 appeared to have frequent signs of liver dysfunctions than mild cases. Increased levels of alanine aminotransferase (ALT), aspartate aminotransferase (AST), and total bilirubin levels are also observed in critically ill patients in ICU^7^. Moreover, administration of various antimicrobials, anti-inflammatory along with antivirals may add some alteration in hepatic biotransformation and biomarkers. Elevated levels of hepato-renal biomarkers are also documented after administration of remdesivir and or favipiravir among the patients admitted in ICU^8^.

Varying clinical features and degrees of severity of COVID-19 leading to multiple approaches to hospital care. It can range from isolation ward to intensive care where patients eventually need to intubate with artificial ventilation^9^. Moreover, demographic and epidemiological characteristics of critically ill patients played a vital role in the duration of stay in ICU, like age or related comorbidities. As this is a novel coronavirus, the length of stay in ICU ranges between ages; even between survivor or non-survivor^10-12^. The need for advanced treatment or intensive care support was higher when most of the ICU was saturated in most of the affected countries including Bangladesh during the peak of the pandemic (May to September 2020). Considering the fact of densely populated 170 million population and low-middle income economy, Bangladesh had constrained of healthcare delivery system to tackle the SARS-CoV-2 endemic. Since the detection of the first case of COVID-19 on 8 March 2020, the complete analytical study of biomarkers as a predictor in critical COVID-19 patients admitted in ICU was never revealed in Bangladesh. This retrospective observational study was therefore done to observe demography, comorbidities, biomarkers, pharmacotherapy, and ICU-stay concerning the mortality outcome of COVID-19 patients admitted in the intensive care unit of the prominent tertiary care hospital to compare between survived and deceased patients.

## Methodology

The cross-sectional retrospective cohort study on critically ill COVID-19 patients admitted to the intensive care unit (ICU) of one of the prime tertiary care hospitals in Dhaka, designated as “COVID-dedicated” for four months by the Government of Bangladesh. All the 112 admitted patients in ICU of Holy Family Red Crescent Medical College Hospital (HFRCMCH) during May to September 2020, designated as the peak of the pandemic were screened and a total of 99 patients were included as per availability of data from the hospital records (non-electronic) and treatment sheets. The HFRCMCH was a 720-bed leading non-government hospital with a 9-bed ICU in the capital city. All consecutive patients with confirmed COVID-19 (by RT-PCR of the nasopharyngeal swab) infection admitted to the ICU in between the time frame were coded critical as per ‘National Guideline on ICU Management of Critically ill COVID-19 Patients (version-8.0)^13^.

Demographic data, comorbidities, duration of ICU-stay, predictive biomarkers (hematological, inflammatory, renal, hepatic, cardiac, and metabolic), and extent of medication used to treat critically ill COVID-19 patients in ICU were collected from hospital records, compiled, analyzed, and compared between improved and dead.

The study was approved by the designated hospital authority and the institutional ethics board (IERB/32/Res/Oct/2020/19/hf). Continuous variables were expressed as the mean and standard deviations; categorical variables were summarized as the counts and percentages and statistical analysis (Chi-square test, unpaired t-test, Fisher’s exact test) was done using SPSS version 26.0 and all *p* values were two-tailed, with *P* <0.05 considered statistically significant with a 95% confidence interval.

## Results

The mean age of the critically ill COVID-19 patients admitted in the ICU was 61.08 years. Among them, 60 patients survived and 39 deceased with the mean age of 57.28 years and 65.02 years, respectively. The majority of the admitted patients were male (72.72%) and the male: female ratio was 1: 2.66 with an age range from 18 to 74 years. Most of the ICU patients were older males within 60 - 69 years of age group (34.34%). The mean age of the patients who survived and deceased was 58.24 (±12.00) and 65.02 (±11.85) years respectively. The overall mortality outcome of critically ill patients in ICU was 39.39%, and sixty patients (60.60%) survived and were discharged alive from ICU. There was a statistically highly significant difference (P <0.005) in mortality across the age groups as shown in Table-1.

**Table-I:**
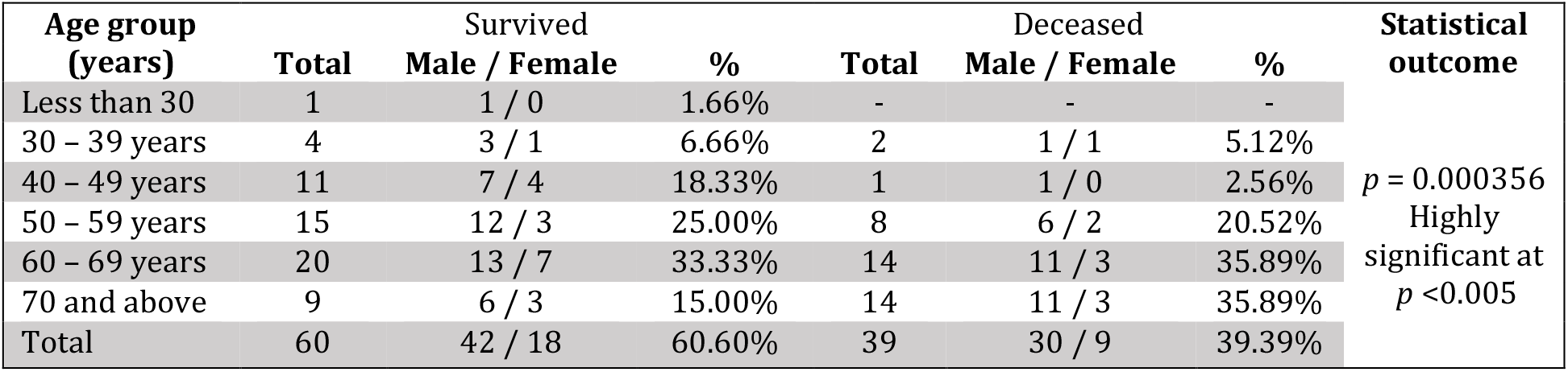
Distribution of patients admitted in ICU according to the age group (*n*=99)

**Table-II:** Biomarkers of critical COVID-19 patients in ICU (*n*=99)

| Biomarkers | Mean ± SD | Total ($n= 99$) | Survived ($n= 60$) | Deceased ($n= 39$) | Statistical value |
| --- | --- | --- | --- | --- | --- |
| *Hematological* | **Total WBC count** | 10,532 ± 4,175 | 9,924 ± 4,386 | 11,382 ± 3,755 | $p = .1156$. |
| | **Neutrophil** | 8569.23 ± 3875.65 | 7807.78 ± 3993.54 | 9635.63 ± 3485.58 | * $p = .0322$ |
| | **Lymphocyte** | 1497.14 ± 1011.03 | 1620.39 ± 1060.18 | 1324.60 ± 925.19 | $p = .1878$ |
| | **NLR** | 7.56 ± 5.57 | 5.99 ± 3.99 | 9.76 ± 6.68 | ** $p = .0017$ |
| | **d- NLR** | 5.68 ± 4.60 | 4.36 ± 2.39 | 7.53 ± 6.13 | ** $p = .0014$ |
| | **Platelet** | 298.41 ± 116.36 | 314.64 ± 130.44 | 273.81 ± 86.62 | $p = .0993$ |
| | **PLR** | 266.91 ± 178.18 | 241.49 ± 146.95 | 305.47 ± 214.08 | $p = .1213$ |
| *Inflammatory* | **Ferritin** | 897.20 ± 644.04 | 841.71 ± 612.28 | 954.19 ± 704.17 | $p = .5457$ |
| | **CRP** | 34.62 ± 27.55 | 36.14 ± 24.62 | 32.13 ± 27.4 | $p = .5652$ |
| | **d- Dimer** | 1.43 ± 2.05 | 1.36 ± 2.12 | 1.57 ± 2.01 | $p = .6652$ |
| *Renal* | **S. Creatinine** | 1.76 ± 1.91 | 1.39 ± 1.08 | 2.303 ± 2.57 | * $p = .0279$ |
| *Hepatic* | **ALT** | 56.34 ± 44.28 | 59.84 ± 50.99 | 64.22 ± 41.56 | $p = .9953$ |
| | **Prothrombin time** | 15.97 ± 2.66 | 15.62 ± 3.53 | 16.27 ± 2.6 | $p = .4783$ |
| | **APTT** | 45.31 ± 9.81 | 43.83 ± 7.4 | 47.57 ± 12.51 | $p = .3433$ |
| | **INR** | 1.20 ± 0.27 | 1.21 ± 0.17 | 1.26 ± 0.20 | $p = .2212$ |
| *Cardiac* | **Troponin- I** | 0.300 ±0.905 | 0.094 ±0.19 | 0.563 ±1.31 | *p* = .0953 |
| *Metabolic* | **Total Cholesterol** | 138.34 ±71.32 | 138.81 ±55.33 | 137.76 ±89.08 | *p* = .9649 |
|  | **Triglyceride** | 225.54 ±94.59 | 147.46 ±75.99 | 187.65 ±97.17 | * *p* = .0432 |
|  | **HDL** | 28.42 ±10.01 | 29.14 ±10.46 | 27.53 ±9.67 | *p* = .6279 |
|  | **LDL** | 73.32 ±41.65 | 80.43 ±45.55 | 64.55 ±35.63 | *p* = .2478 |
|  | **HbA<sub>1</sub>C** | 7.45 ±1.04 | 7.36 ±1.04 | 7.56 ±1.06 | *p* = .5676 |

Pre-existing hypertension (64.64%) and diabetes mellitus (71.71%) were common among both the survived and deceased patients. Ischemic heart disease was also found in 21.21% of patients in ICU. But COPD (15.38%) and chronic kidney diseases (30.76%) were much higher among the deceased cases. Other comorbidities (hyperlipidemia, carcinoma, thyroid dysfunction, bronchial asthma) were also higher (28.20%) in deceased patients (Figure-1). But there was no statistical significance of difference (*p* = 0.232) between survived and a deceased group of patients.

**Figure-1:**
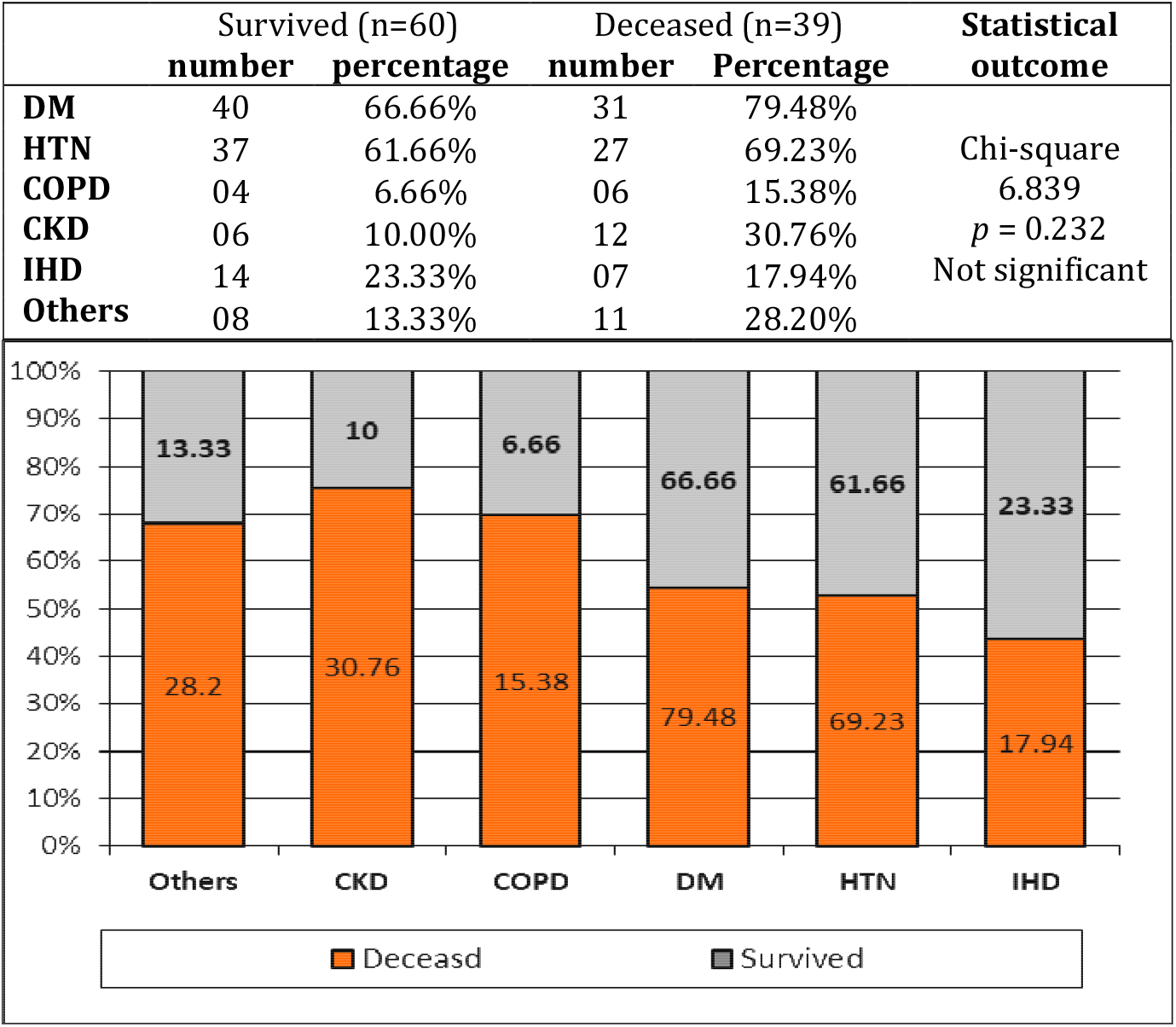
Comorbidities among survivors and non-survivors in ICU

The duration of stay in ICU was observed for all 99 critically ill COVID-19 patients during the study period of four months. Most of the critical patients (25) died within 5 days of stay in ICU while survived only 17 patients. But the mortality rate was linearly decreased with longer duration of ICU stay while survival rate was much higher at 10-day stay (8 vs 13), 20-days stay (5 vs 23) and 30-days stay (1 vs 7) in ICU. Most of the patients survived who needed to stay up to 20 days in ICU to manage critical complications (Figure-2)

**Figure-2:**
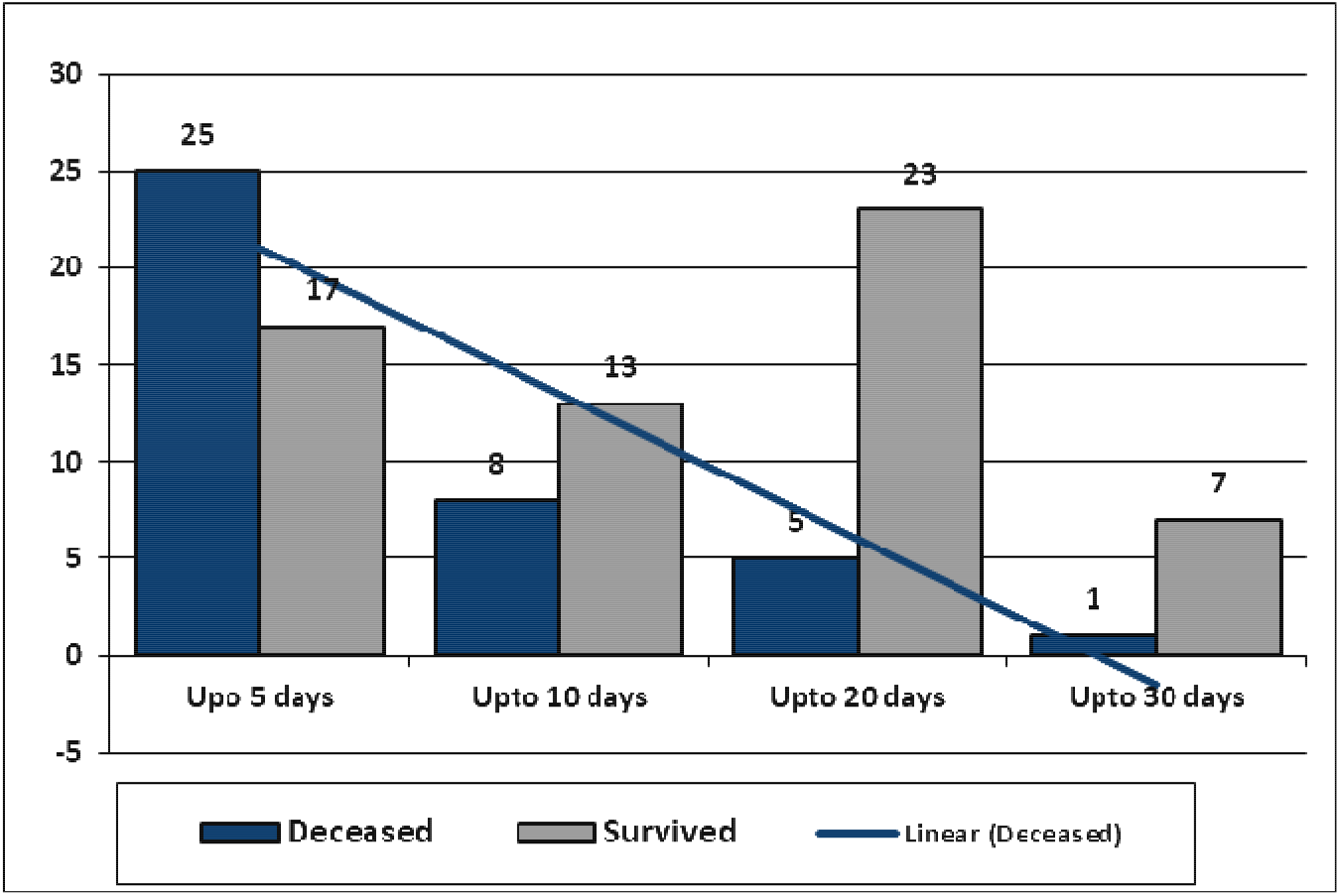
Mortality trend according to the duration of ICU-stay

To compare the statistical significance of variation between survived and deceased patients, an unpaired *t*-test was done for every predictor, and the difference between two outcome groups, NLR (neutrophil-lymphocyte ratio) and d-NLR was highly significant (*p* =.0017 and .0014). The total count of neutrophils was also significantly increased (*p* =.032) among the deceased patients. Differences in inflammatory and metabolic biomarkers were non-significant except for the serum triglyceride (*p* =.0432). Hepatic and cardiac markers were also increased among deceased patients but were not significant statistically. But serum creatinine was significantly (*p* =.0279) high among deceased patients than the survived as 2.303 ±2.57 and 1.39 ±1.08 respectively.

Besides wide variation of pharmacotherapy, the use of meropenem was the most common drug used among survived (93%) and deceased (92.30%) patients treated in ICU. Moxifloxacin was the second most frequently used antibiotic in 90% and 84.61% of patients respectively. Almost 88.33% of survived patients and 82.05% of deceased patients received remdesivir.

Moreover, favipiravir, tocilizumab, and cephalosporin were also used. There was no statistical significance of difference as mortality outcome by chi-square (Figure-3).

**Figure-3:**
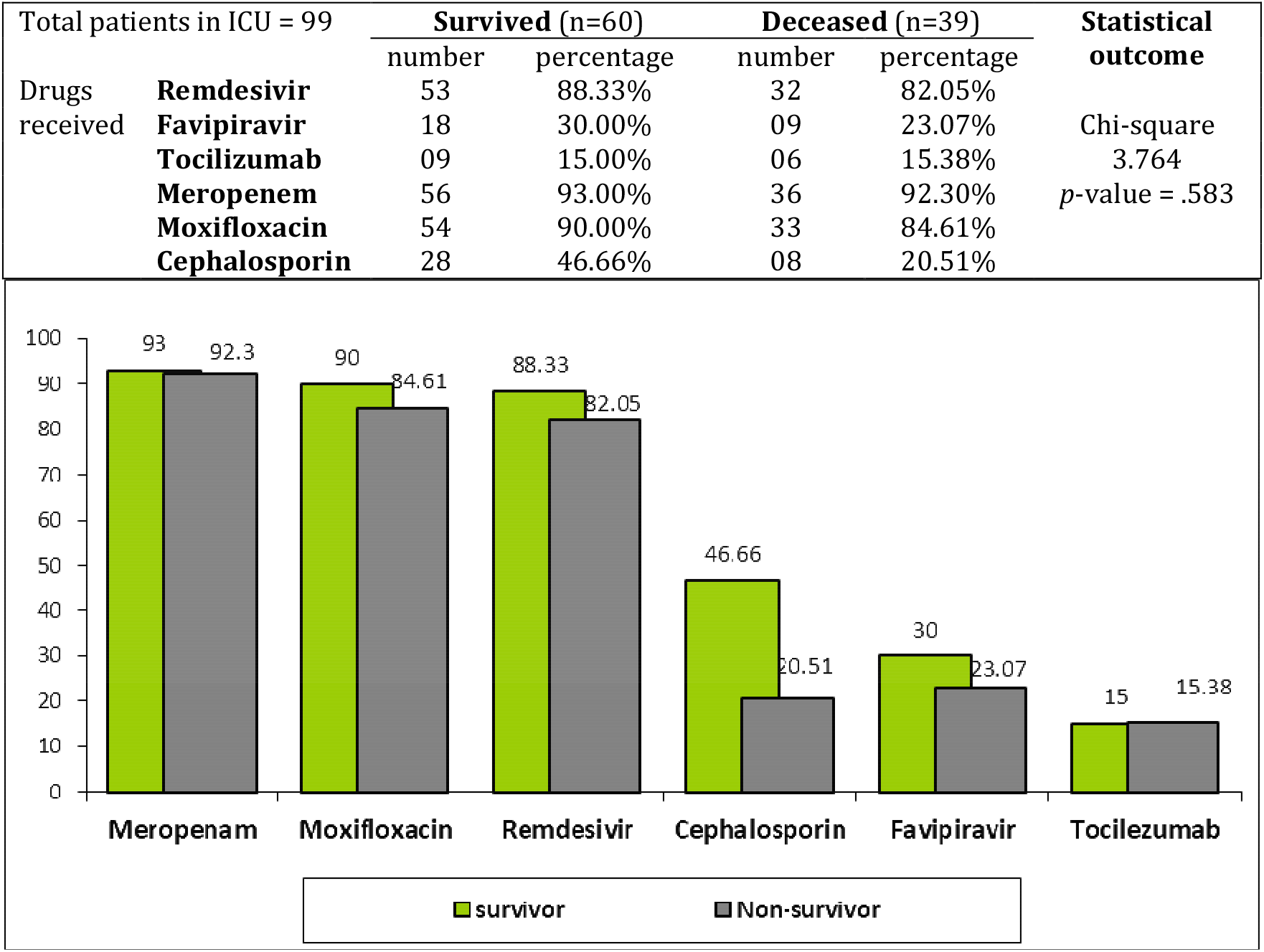
Drugs used in ICU among survivors and non-survivors

## Discussion

Coronavirus Disease 2019 (COVID-19) is an ongoing global pandemic causing significant increases in morbidity and mortality, whose clinical impact is particularly severe for older individuals having frequent serious comorbidities act as relevant predisposing factors for a more severe COVID-19 clinical course and consequent death^14^. Such high mortality is prominently due to severe acute respiratory syndrome, able to quickly spread to vulnerable populations such as comorbid elderly individuals with aging-related disorders. The number of male patients admitted in ICU with the critical condition was much higher than the female (1: 2.66) and the survival rate was also proportionately higher in males, which was similar to the other studies around the globe. In Bangladesh, several other studies on the patients admitted in ICU also revealed 77.5%, 64.28%, 77% in Bangladesh^6,8,15^ and 66.30% in Mexico^16^, almost 65% in the USA^17^ accordingly. The mean age of the patients who survived (57.28) and deceased (65.02) were also found to be similar among the critical patients in other studies, and most of the admitted patients were above 60 years of age^6,8,18^.

Pre-existing conditions such as chronic kidney disease (CKD), cardiovascular disease, chronic lung disease (particularly COPD), diabetes mellitus (DM), hypertension (HTN), immunosuppression, and obesity evolved the COVID-19 patients into critical courses, leading to increased mortality with mechanical ventilation in different countries^10,19,20,21^. However, almost two-thirds of the patients in ICU having CKD were deceased in our study cohort whereas DM and HTN were the most common comorbidities in both survivors and a deceased group of patients. A similar finding was observed in other studies conducted in hospitals in Dhaka, Bangladesh^22,23^. Whereas, pre-existing COPD and ischemic heart diseases were reported as more common comorbidities among critically ill COVID-19 patients in ICU of the UK, USA, and Italy. In China, these comorbidities include diabetes (7.3%), respiratory disease (6.5%), cardiovascular disease (10.5%), hypertension (6%), and oncological complications (5.6%) and patients without comorbidities have a lower case fatality rate (0.9%)^24^. These variations are yet to be observed more keenly among larger-scale epidemiological studies.

In the present study, the duration of stay in ICU was inversely proportional to the mortality rate. Almost 59.52% of critically ill patients died within 5 days of disease progression. However, the mortality rate was much declined in 10-days, 20-days, and 30-days of ICU stay as 38%, 17.8%, and 12.5% respectively. This might be due to the management of pre-existing diseases and continuous clinical assessment. The overall mortality rate was 39.39% in four months which was almost similar to the outcome in China (37.7%), Denmark (41.2%), and the United States (38.6%) at ICU of tertiary care hospitals^25,26^. Another pilot study on 58 critically ill patients in ICU who received remdesivir or favipiravir has reported a higher mortality rate of 44.8% among the similar cohort in Bangladesh^27^.

The most unpredictable predictors of the COVID-19 patients range from hematological markers to inflammatory and immunological biomarkers until today. In the present study, most of the biomarkers of Covid-19 patients admitted in ICU were observed and compared between the survived versus deceased groups. A significant rise of neutrophil (p< .032), NLR (neutrophil-lymphocyte ratio) (p< .0017), and d-NLR (p< .0014) was found in our cohort but raised total count of WBC and PLR (platelet-lymphocyte ratio) was not statistically remarkable with the death group. Lymphopenia and thrombocytopenia were also found in almost all the critically ill COVID-19 patients in our study. However, these findings were variable with the studies done in Seattle, New York City, Chicago, and Washington States^28,29^ where the lymphopenia was common but the neutrophil count was not elevated among the patients in ICU. Moreover, the significant increase of NLR and d-NLR was reported to be associated with an 8% increased risk of mortality in China^30^. Qin *et al* also reported a significantly higher NLR (in a cohort of 452 severe COVID-19 patients^31^ and prognostic role of PLR by Qu et al^32^. However, the present study showed similarity with NLR but dissimilarity with the PLR as a dependable predictor among the deceased group of patients. A report on 74 COVID-19 patients from the Italian frontline predicted clinical improvement with an NLR <3 and critically required admission to ICU with an NLR >4 which corresponds to our study^33^. Among the inflammatory biomarkers, CRP, d-Dimer, and ferritin were high among the non-survivors in different studies and reported as a significant predictor for severity of illness^28,34^. But no significant differences were observed between the survivors and deceased patients in the present study. Besides following the standard guideline of the treatment protocol, this might be due to racial and demographic variation of population arising further question about the consistency of predictors for COVID-19. A significant difference of increased creatinine and triglyceride was found between the survived and dead patients in our study, whereas ALT, PT, APTT, and INR were non-significant that differs from other studies as a predictor of hypercoagulability^35^. The overall elevation of inflammatory, hepatic (ALT), and cardiac (Troponin-I) biomarkers were reported in studies in the US population28 but were non-significant in our cohort as a predictor of mortality outcome among critically ill COVID-19 patients in ICU. Changes in the metabolic (HbA1C, lipid profile) biomarkers were also unremarkable.

The current ongoing pandemic of COVID-19, together with a huge effort to discover new drugs or vaccines able to stop the virus spreading, also requires repurposing of an existing drug as a safe and effective alternative potentially able to fight the critical conditions2. Besides the repurposing use of antivirals, the present observational study revealed that meropenem was the most commonly used antimicrobial in almost all the critical patients in ICU, followed by moxifloxacin. Use of remdesevir (88.33% vs 82.05%) and favipiravir (30% vs 23.07%) showed no significant difference in mortality outcome between survivors and deceased groups of patients in ICU. Almost similar findings were also observed in a previous study in Bangladesh6. The chances of respiratory infection by nosocomial and opportunistic microbes might be a critical concern for patients admitted in ICU in Bangladesh with poor ICP (infection control and prevention) that justify the use of a wide variety of antibiotics including cephalosporin in the present study.

## Conclusion

The long-standing pandemic of SARS-CoV-2 coronavirus infection is presenting with different strains, clinical impressions, and variations of predictive biomarkers with geographical diversity. The predictors of severity, progression, and high mortality of critically ill COVID-19 patients in ICU require empiric observation and likely to evolve rapidly in different populations. Therefore, the global focus should be to keep pace in search for consistent, early predictors of disease progression for effective treatment and management of critical care facilities to save COVID-19 patients in ICU, particularly in a low-middle income country like Bangladesh.

## Data Availability

All data are available on request

